# Polygenic Risk Factors for Comorbid Diagnoses in Individuals with Substance Use Disorders: A Phenome-Wide Survival Analysis

**DOI:** 10.1101/2024.12.13.24319000

**Authors:** Peter B. Barr, Zoe E. Neale, Tim B. Bigdeli, Chris Chatzinakos, Philip D. Harvey, Roseann E. Peterson, Jacquelyn L. Meyers

## Abstract

**Objective:** Persons with substance use disorders (SUD) often suffer from additional comorbidities. Researchers have explored this overlap via phenome wide association studies (PheWAS). However, PheWAS are largely cross-sectional, limiting our understanding of whether diagnoses predate development of an SUD. We characterize whether polygenic scores (PGS) are associated with time to comorbid diagnoses in electronic health records (EHR) after the first documented SUD diagnosis.

**Methods:** Using data from All of Us (N = 393,596), we explored: 1) whether social determinants of health (SDoH) are associated with lifetime risk of SUD (N cases = 42,568) and 2) within a subset those with a diagnosed SUD and available genetic data SUD (N = 21,357), whether PGS for alcohol use disorders, cannabis use disorders, depression, externalizing, post-traumatic stress disorder, and schizophrenia were associated with subsequent diagnoses via a phenome-wide survival analysis.

**Results:** Multiple SDoH were associated with lifetime SUD diagnosis, with annual household income having the largest overall associations (e.g., <$10K annually vs $100K-$150K annually: OR = 4.18; 95% CI = 3.92, 4.45). There were 86 phenome-wide significant PGS associations with subsequent diagnoses across various bodily systems. PGSs for alcohol use disorders, post-traumatic stress disorder, and schizophrenia were each associated with time to their respective diagnoses.

**Conclusions:** Social determinants, especially those related to income, have profound associations with lifetime SUD risk. Additionally, PGS for psychiatric conditions are associated with multiple post-SUD diagnoses within those with a SUD, suggesting PGS may capture information beyond lifetime risk, including timing and severity of comorbidities related to SUD.

## INTRODUCTION

Psychiatric disorders have far-reaching consequences for affected individuals, their families, the communities in which they live, and society more broadly (1–4). Substance use disorders (SUD) in particular are extremely detrimental. An estimated 107,000 Americans died as the result of an overdose in 2021 (5). In 2016, alcohol use contributed 4.2% to the global disease burden and other drug use contributed 1.3% (4). Excessive alcohol use and illicit drug use cost the United States an annual $250 billion (6) and $190 billion (7) respectively. Part of the increased mortality and morbidity in those with SUD occurs because if the increased number of co-occurring medical problems associated with having a SUD. Given the substantial human and economic costs of substance use disorders, understanding the ways SUD can contribute to risk for comorbidity has important public health implications.

The proliferation of large-scale biobanks – such as The Million Veteran Program (8), FinnGen (9), Biobank Japan (10), UK Biobank (11), and more recently, All of Us (12) – linking individual-level genomic data with electronic health records (EHRs) presents opportunities to further explore the relationships between SUD, or any contributory indicators (e.g., clinical diagnoses, polygenic scores), and a wide range of outcomes. This hypothesis-free approach, referred to as a phenome-wide association study (PheWAS) (13), can aid us in better understanding patterns of comorbidity with any given exposure. For example, recent PheWAS using polygenic scores (PGS) for schizophrenia, bipolar disorder, major depression, and broad externalizing risk (14–16) have identified widespread associations between PGSs and a host of psychiatric and other medical diagnoses spanning all bodily systems and have significant potential implications for public health strategies.

To date, PheWAS have primarily used cross-sectional data. This lifetime-only approach limits the potential to understand: 1) whether certain “exposures” occur after the onset of a given condition, obscuring the time-order of comorbid diagnoses, and 2) the degree to which diagnoses may lead to a cascade of other comorbidities. For example, using lifetime smoking as the exposure, we could potentially misinterpret comorbid psychiatric conditions as consequences of smoking in the case when they are a risk factor for smoking. Additionally, we may miss the consequences of heavy smoking, and time to their onset, if we do not focus on smoking quantity and post-initiation diagnoses among smokers. In the current study, we move beyond lifetime diagnosis in PheWAS to characterize whether different PGS are associated with time to comorbid diagnoses in after the first indication of SUD in EHRs. We do so by utilizing longitudinal EHR data from the All of Us Research Program, which is more demographically representative than previous work in this area.

The clinical utility of PGSs is an ongoing area of debate (17–22). We add to this discussion in three ways. First, we explore the potential use of polygenic scores *within individuals who are already diagnosed*. We chose substance use disorders specifically because of the wide array of medical and psychiatric consequences related to having a SUD. Second, we move beyond a focus of lifetime associations in PheWAS to characterize whether polygenic scores are associated with subsequent comorbid diagnoses appearing in the EHR *after the first recorded entry of substance use disorder* (SUD). Exploring the post-SUD diagnosis association of polygenic scores (PGS) for psychiatric and substance use disorders with subsequent medical diagnoses can help characterize the vast pleiotropic impact of genetic risk for these conditions, which could reflect shared genetic effects (biological pleiotropy, e.g., a common liability contributes to risk for both SUD and post-traumatic stress disorder, or PTSD, explaining the comorbidity) or a potential causal chain of events (mediated pleiotropy, e.g., liability for AUD contributes to increased alcohol use, resulting in liver disease). Lastly, we harness the diversity in All of Us to move beyond the limited focus on populations most similar to European reference panels. Knowledge gained regarding the potential medical complications likely to arise in individuals with SUD with and without genetic risk for SUD can lead to preventative actions or improved screening that may reduce the onset of these additional comorbidities.

Using data from the All of Us Research Programs’ EHR (Release 8, N = 393,596), we first explored whether several social determinants of health were associated with lifetime risk of SUD, given their importance in the onset of SUD (23).We did this, in part, to characterize patterns of diagnosis and assess potential biases when selecting those with a SUD in EHR. Conditioning on those that have a diagnosis increases the potential for collider bias in downstream analyses (24), as seen in recent analyses of polygenic scores within those that have psychotic disorders (25). Next, harnessing the longitudinal quality of All of Us, we examined whether PGSs for six psychiatric and substance use disorders were associated with subsequent comorbidities post-SUD diagnosis, by performing a phenome-wide survival analysis. Our analyses inform the degree to which genetic and environmental risk factors are important in the course of SUD and related medical problems. While the PheWAS is exploratory in nature, we expect: 1) polygenic scores for a psychiatric conditions to be associated with subsequent diagnoses for their respective phenotype, and 2) these polygenic scores to be associated with subsequent comorbidities relevant for each condition (e.g., AUD PGS and liver conditions).

## METHODS

### The All of Us Research Program

All of Us is a prospective, nationwide cohort study aiming to study the effects of environment, lifestyle, and genomics on health outcomes. Participant recruitment is predominantly done through participating health care provider organizations and in partnership with Federally Qualified Health Centers. Interested participants can enroll as direct volunteers, visiting community-based enrollment sites. Enrollment, informed consent, and baseline health surveys are administrated digitally through the All of Us program website (https://joinallofus.org). Participants are next invited to undergo a basic physical exam and biospecimen collection at an affiliated healthcare site. Participant follow-up occurs through both passive linkage with EHR and active periodic follow-up surveys. We included the full sample with EHR data for associations between social risk factors and lifetime SUD diagnosis (N = 393,596). For the genomic analyses, we included all participants who had electronic health record (EHR), had short-read whole genome sequence (WGS) data from release 8 (May 6, 2018 to October 1, 2023), and whose genetic similarity matched populations from available genome wide association studies (GWAS). Additionally, we excluded those with self-reported military service to remove potential overlap with the GWASs used to generate PGS, such as GWAS from the Million Veteran Program, leaving the final sample with a lifetime SUD and genomic data at *N =* 21,357.

### Genotyping

In the current release of All of Us (release 8), N = 414,840 participants have available short-read sequencing data. To ensure consistency across sites for DNA extraction and sequencing, AoU developed a standardized process and QC metrics across sites. Sequencing was performed on the Illumina NovaSeq 6000 instrument. A full description of the collection, harmonization, sequencing, and population assignment pipeline has been published elsewhere (26). To correct for population structure in the genetic data, participants were assigned to genetic similarity clusters of those most similar to European (EUR-like) and African (AFR-like) participants in the 1000 Genomes and the Human Genome Diversity Project (27–29). Genetic principal components (PCs) were generated in Hail (30).

### Electronic health records (EHRs)

We used phecodes, which are validated clusters of ICD-9/10-CM codes in the EHR (31–34), to create lifetime diagnoses for SUD (i.e., across the entire course of one’s EHR) and the subsequent outcomes for the PheWAS. We considered individuals as meeting criteria for an SUD if they had two or more outpatient or one inpatient occurrence of phecodes 316 (Substance addiction and disorders, tobacco-related disorders excluded) or 317 (Alcohol-related disorders) in their EHR. Prior analyses have shown that 2 or more phecodes is a good predictor of diagnosis, and we chose this definition to limit diagnostic bias (14, 15). The date of the first recorded SUD in participants EHR was used as the index date for all longitudinal analyses. We focus on diagnoses of alcohol and drug-related disorders, excluding those *only* diagnosed with tobacco use disorders. Alcohol and other drug use disorders have substantial genetic overlap (35, 36) that overlaps relatively less with risk for tobacco related disorders (37–39). We performed a series of sensitivity analyses including tobacco use disorders (phecode 318) to examine whether the inclusion of tobacco use disorder changed the pattern of results. For PheWAS outcomes, we used the presence of any phecode following the initial diagnosis of SUD. We excluded phecodes that occurred at a rate lower than 1% in each population. Our choice to use a single phecode following SUD was driven by: 1) the need to ensure adequate power given the smaller overall sample size for SUD cases and relative infrequency of some EHR codes, and 2) many of the common comorbidities (e.g. obesity, hypertension, type 2 diabetes) lack the level of diagnostic uncertainty present in psychiatric diagnoses that more often require a higher threshold for certainty.

### Social and demographic risk factors

We included a variety of social and demographic measures available from the baseline survey. For the current analysis we included age, gender (man, woman, or trans and gender nonconforming), self-reported race-ethnicity (Asian American/Pacific Islander, Black/African-American, Hispanic or Latino/a/x, Multi-racial, Non-Hispanic White, and other race-ethnicity), education (less than high school, HS diploma or GED, some college, college graduate, and advanced degree), household income (less than $10k to more than $200), marital status (never married, married, cohabitating, divorced, separated, and widowed), health insurance (insured vs uninsured), and place of birth (US born vs foreign born). Our reference categories (married, Non-Hispanic White, college graduate, etc.) were selected because they reflect those that are typically most socially advantaged and have better health, on average. We focused on the association between these social risk factors and lifetime SUD diagnosis, only. Because these items are measured at baseline entry into All of Us and not necessarily time of SUD diagnoses, we lack the ability to establish whether potentially time-variant measures were the same before their first SUD diagnosis.

### Polygenic scores (PGS)

We estimated PGSs, which are aggregate measures of the number of risk alleles individuals carry weighted by effect sizes from GWAS summary statistics, from multiple large-scale GWASs for alcohol use disorders/problematic alcohol use, cannabis use disorders, broad externalizing risk, depression, post-traumatic stress disorder, and schizophrenia (40–46). We included these specific PGSs because: 1) there is strong genetic overlap between psychiatric conditions disorders, 2) these PGS cover a range of risk for internalizing, externalizing, and thought disorders (40–42, 44, 47, 48), and 3) each of these GWAS included results for AFR-like participants, allowing us to move beyond EUR-like only analyses. We created PGSs using PRS-CSx (49), a Bayesian regression and continuous shrinkage method that estimates the posterior effect sizes for each SNP in a given set of GWAS summary statistics. PGS accuracy decays continuously as target samples differ in ancestral background from the discovery GWAS, even within relatively homogenous genetic clusters (50). PRS-CSx uses multiple populations to improve power of PGS in underpowered samples, typically those who are not in the EUR-like grouping. For PRS-CSx, we used the *auto* flag to identify the optimal shrinkage parameter (phi), the *meta* flag to generate the weighted effect sizes estimates, and the default settings for all other flags. We standardized all PGSs to Z-scores for interpretation.

We note that our paper includes language related to both race-ethnicity, which reflects socially-constructed categories, and genetic similarity, which uses empirical assignment based on available reference panels, because both are relevant for the current analyses. Prior work has established race-ethnicity (and with it, racism and discrimination) are relevant for disparities in SUDs (51–55). Informed by best practices for handling genetic data from diverse populations (56), we stratified analyses by genetic similarity and included genetic principal components, which limit the possibility of false positives due to population stratification. Race-ethnicity was used in the analyses related to sociodemographic risk factors while genetic similarity was used in PGS analyses. The inclusion of both concepts is in no way endorsing the notion that these reflect discrete biological categories.

### Electronic Health Record (EHR) specific covariates

We included several covariates specific for EHR-based analyses. First we construct measures for length of EHR (time from first to most recent documented code) and healthcare utilization (total count of documented billing codes across the EHR record) using the *PheTK* package, specifically designed for work in All of Us (57). To account for comorbidities that predated SUD diagnosis, we created comorbidity burden score by taking the inverse normal transformation of the total count of unique phecodes individuals had in their EHR prior to SUD, used in previous PheWAS analyses (15, 16).

### Analytic plan

Figure 1 demonstrates how the analytic samples for each of the analyses were selected. First, we examined the association between social and demographic measures and lifetime SUD diagnosis using logistic regression (N = 393,596). Next, we expanded on the traditional PheWAS approach by performing a phenome wide survival analysis (N = 21,357). We utilized Cox proportional hazards models to estimate the association (in the form of hazard ratios, or HR) between the index date (time of initial SUD diagnosis in EHR) to first documented diagnosis for each phecode, as PheWAS is a hypothesis-free approach which examines *all* outcomes instead of just theoretically relevant ones. The bottom panel in Figure 1 provides a visual depiction of how the timelines might vary for respondents in the survival models. In the PGS models, we included all 6 PGS simultaneously. We stratified by genetic similarity grouping (AFR-like and EUR-like) and included age at SUD diagnosis, gender, the first 10 genetic principal components, pre-SUD diagnosis comorbidity burden, length of EHR, and healthcare utilization as covariates. We did not impose any predefined exclusionary criteria regarding length of EHR record since we used the date of first SUD record as index date and the Cox proportional hazards model is well-suited to handle right censored data. We performed all survival models using the *survival* package (version 3.7.0) in R (version 4.4.1). Finally, we meta-analyzed results across genetic-similarity via fixed-effects meta-analysis in the *meta* package (version 7.0.0) in R. To correct for multiple testing, we applied a false discovery rate (FDR) of 5% (58).

**Figure 1:**
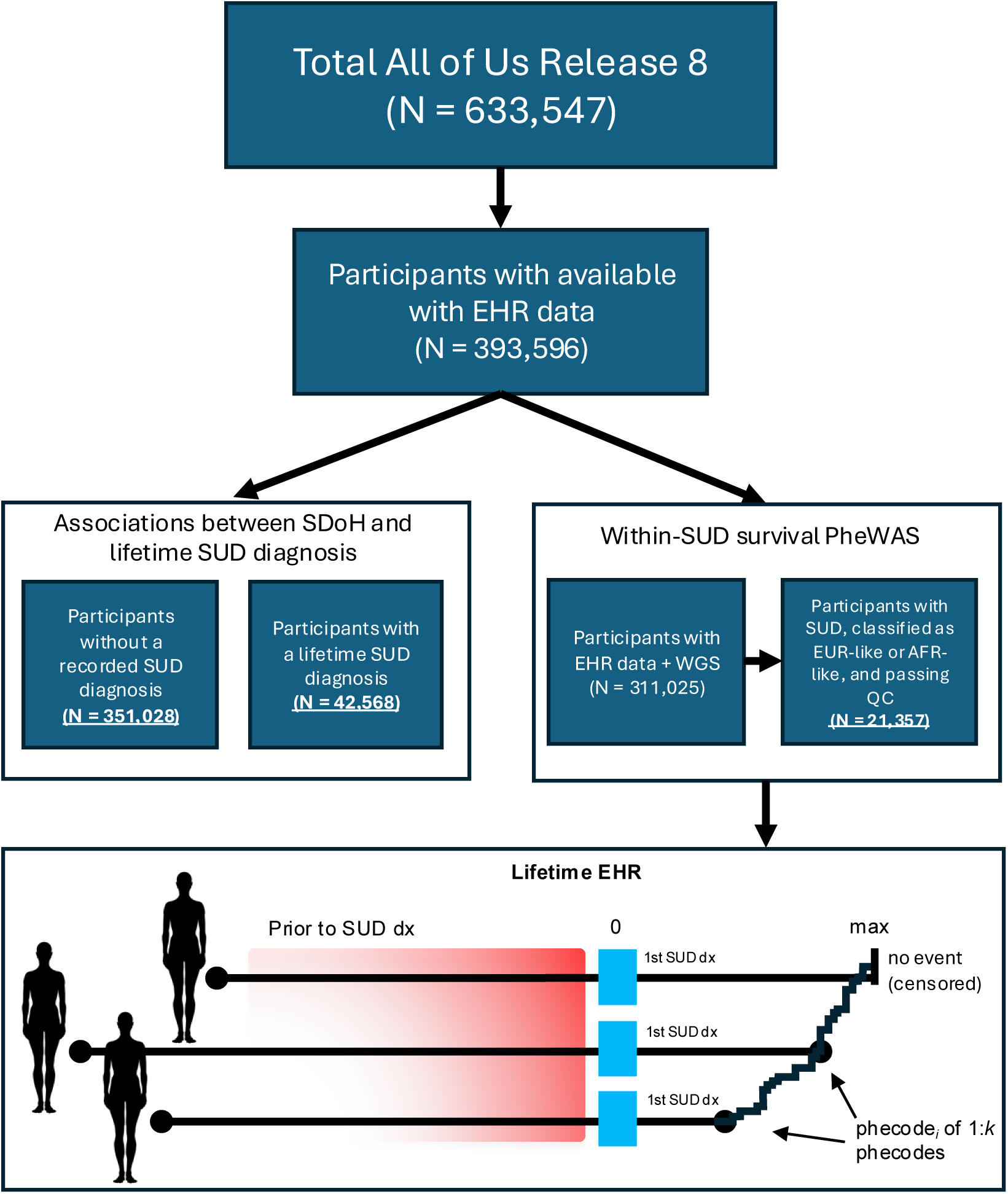
Sample selection in the All of Us Database. Sample selection for analyses of lifetime risk for substance use disorder (N = 393,596) and within-SUD, survival PheWAS (N = 21,357). EHR = electronic health records; WGS = whole genome sequence, QC = quality control.

## RESULTS

### Defining persons with lifetime SUD in EHR

Using the 2+ outpatient/1+ inpatient definition of substance use disorder diagnosis, we identified N = 42,568 individuals who met our criteria for SUD in the full data with EHR available. Table 2 presents demographic characteristics for the full sample with available EHR data and those who met our criteria for SUD. This demographic breakdown of the SUD sample largely reflects the patterns seen in nationally representative samples (51, 52).

**Table 1:**
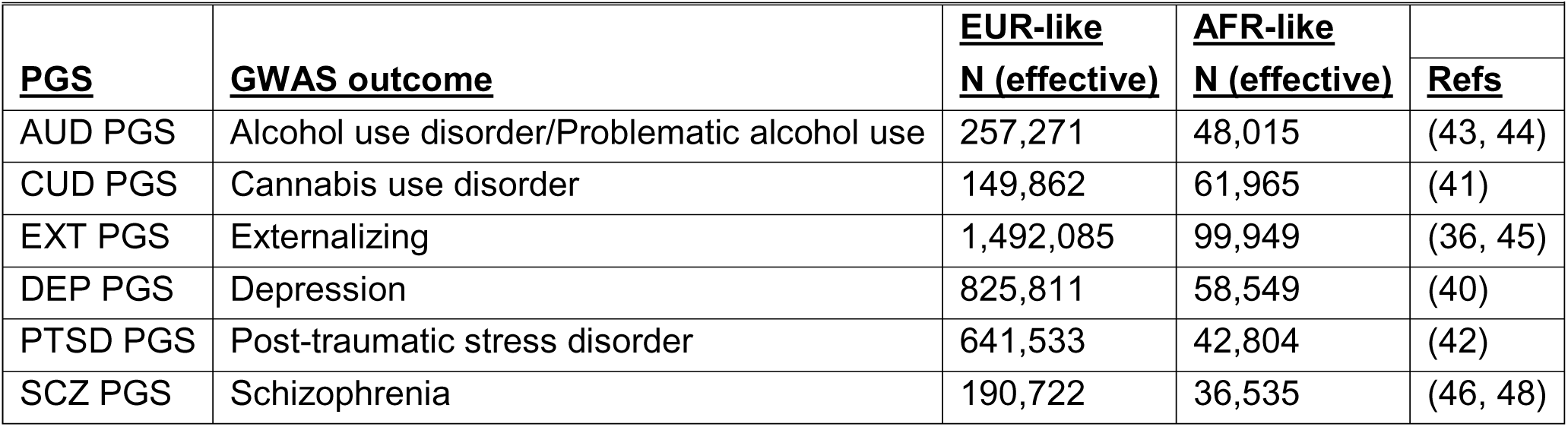
GWAS used input for polygenic score creation.

**Table 2:** Sample Demographics in All of Us participants with available EHR data and those with a SUD diagnosis.

|  | Full sample<br>(N = 393,596) |  | SUD sample<br>(N = 42,568) |  |  |
| --- | --- | --- | --- | --- | --- |
|  | <u>N/Mean</u> | <u>%/SD</u> | <u>N/Mean</u> | <u>%/SD</u> | <u>$\chi^2$</u> |
| Age (mean) | 52.36 | 16.99 | 50.06 | 14.63 | * |
| Woman | 237,501 | 60.34% | 19,501 | 45.81% | * |
| Man | 148,918 | 37.84% | 22,009 | 51.70% | * |
| Trans and gender non-conforming (TGNC) | 3,630 | 0.92% | 434 | 1.02% |  |
| Asian-American/Pacific Islander | 11,714 | 2.98% | 320 | 0.75% | * |
| Black/African American | 67,197 | 17.07% | 11,769 | 27.65% | * |
| Hispanic or Latino/a/x | 59,606 | 15.14% | 5,308 | 12.47% | * |
| Non-Hispanic White | 210,276 | 53.42% | 18,448 | 43.34% | * |
| Multiple race-ethnicities | 27,177 | 6.90% | 3,667 | 8.61% | * |
| Other race-ethnicity not listed | 6,134 | 1.56% | 712 | 1.67% | * |
| Born outside the US | 56,684 | 14.40% | 2,536 | 5.96% | * |
| US-born | 333,579 | 91.89% | 39,414 | 92.59% | * |
| Less than high school | 34,773 | 8.83% | 7,036 | 16.53% | * |
| High school diploma or GED | 74,172 | 18.84% | 13,254 | 31.14% | * |
| Some college | 102,999 | 26.17% | 12,303 | 28.90% | * |
| College graduate | 89,340 | 22.70% | 4,918 | 11.55% | * |
| Advanced degree | 83,228 | 21.15% | 3,402 | 7.99% | * |

\* $p < .05$ ,

### Social and Demographic results

Figure 2 presents the bivariate and adjusted (multivariable) results from the logistic regression models examining the association between social and demographic risk factors and SUD diagnosis in the full sample with available EHR data for two selected SDoH: annual household income and marital status (N = 393,596). These results largely mimicked prior analyses in earlier versions of the All of Us data (59), presented here as odds ratios (OR) and corresponding 95% confidence intervals (CI). There was a stark gradient in lifetime SUD diagnosis across income, whereby those with the lowest income had the greatest odds of lifetime diagnosis (adjusted OR = 4.18; CI = 3.92, 4.45) relative to those reporting annual incomes of $100K - $150K. Additionally those that were separated (adjusted OR = 2.32; CI = 2.20, 2.45) or divorced (adjusted OR = 1.97; CI = 1.91, 2.05) had significantly higher odds of SUD diagnosis relative to those that were married. Results were slightly attenuated when moving from the bivariate to adjusted (inclusion of all social risk factors and covariates simultaneously), but the patterns remained unchanged. The full results are available in Supplemental Tables 1 & 2.

**Figure 2:**
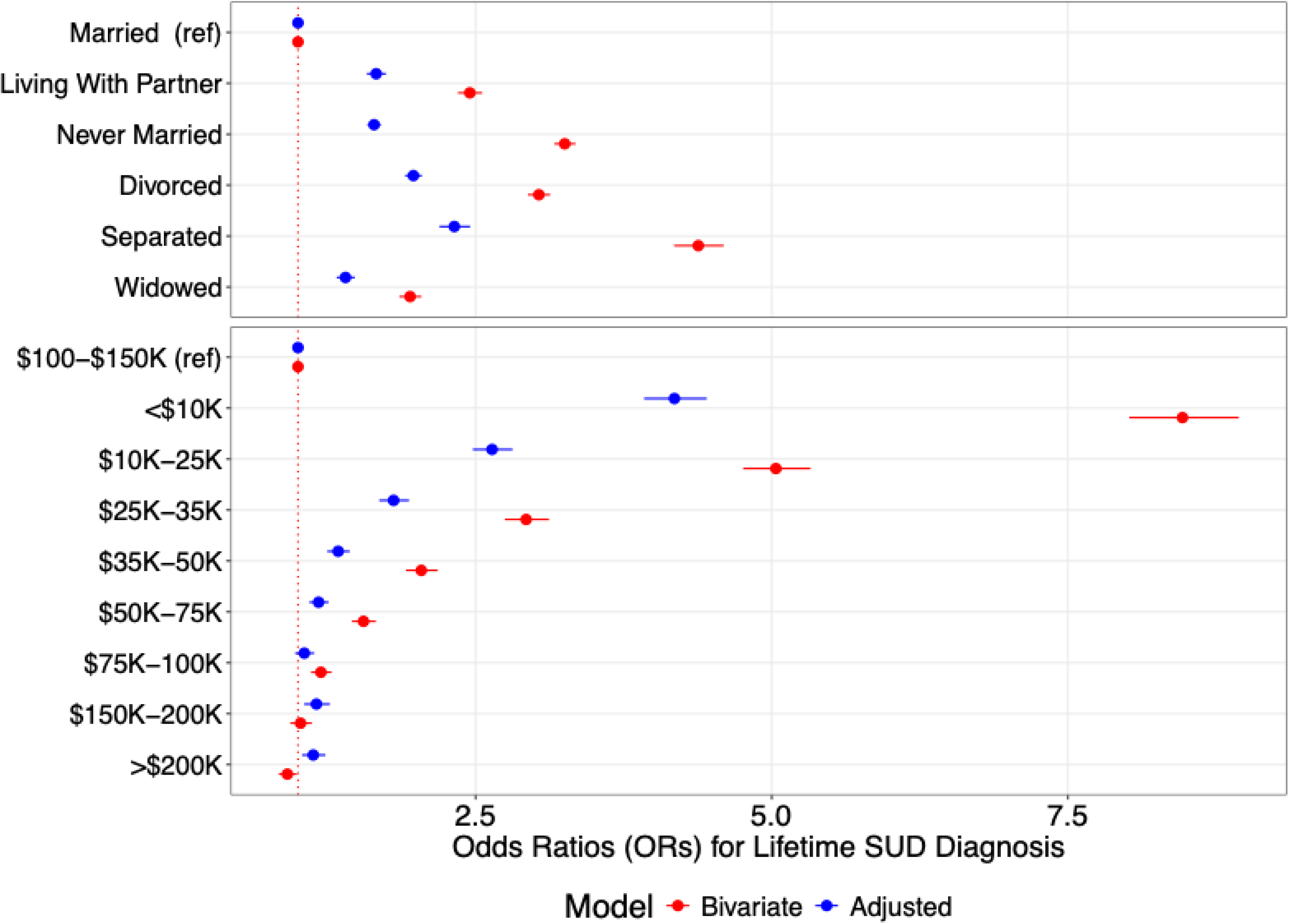
SUD rates across sociodemographic characteristics and top associations from multivariable logistic regression. Bivariate and Adjusted odds ratios (OR) and corresponding 95% confidence intervals presented on the x-axis. Reference categories indicated by (ref). Adjusted models included all of the social and demographic risk factors listed in the methods section, simultaneously. Adjusted models also included covariates for length of EHR and total healthcare utilization.

### Polygenic Score (PGS) Survival PheWAS

Moving to those with an SUD diagnosis only, we further constrained to those of AFR-like and EUR-like genetic similarity for subsequent within-SUD survival models using PGSs (Combined N = 21,357, see Supplemental Table S3 for stratified results). The subset of EUR-like and AFR-like participants did not differ significantly in age from the full SUD sample (mean age = 50.45, SD = 14.24), but there was a significantly higher proportion of women compared to the broader SUD sample (51.22%, χ^2^ = 167.33, df = 1, *p* = 2.83×10^-38^). The vast majority identified as Non-Hispanic White or Black/African American (86.93%). In total, we examined the associations with 846 phecodes following initial SUD diagnosis after filtering those that did not occur in 1% or more of either population (full list of phecodes in Supplemental Table S4). After adjusting for multiple testing, only 86 (10.1%) of the associations remained significant. The proportion that had a given code prior to SUD diagnosis ranged from 0.17% (phecode 797.1, cardiogenic shock) to 31.25% (phecode 745.0, pain in joint, full results Supplemental Table S4), showing that many of these post-SUD codes reflect new incident cases. A selection of the top associations for each PGS are presented in Table 3 (see Supplemental Table S4 for full results). Of all the PGSs, the PGS for schizophrenia (SCZ) showed the greatest number of trait associations, followed by the PGSs for PTSD, externalizing (EXT), and depression (DEP). The PGSs for alcohol use disorder (AUD) and cannabis use disorder (CUD) were each only associated with one future phecode. Importantly, PGSs for SCZ, PTSD, and AUD were associated with time to SCZ (HR = 1.21; 95% CI = 1.15, 1.27), PTSD (HR = 1.13; 95% CI = 1.08, 1.19), and AUD (HR = 1.08; 95% CI = 1.05, 1.11), respectively.

**Table 3:**
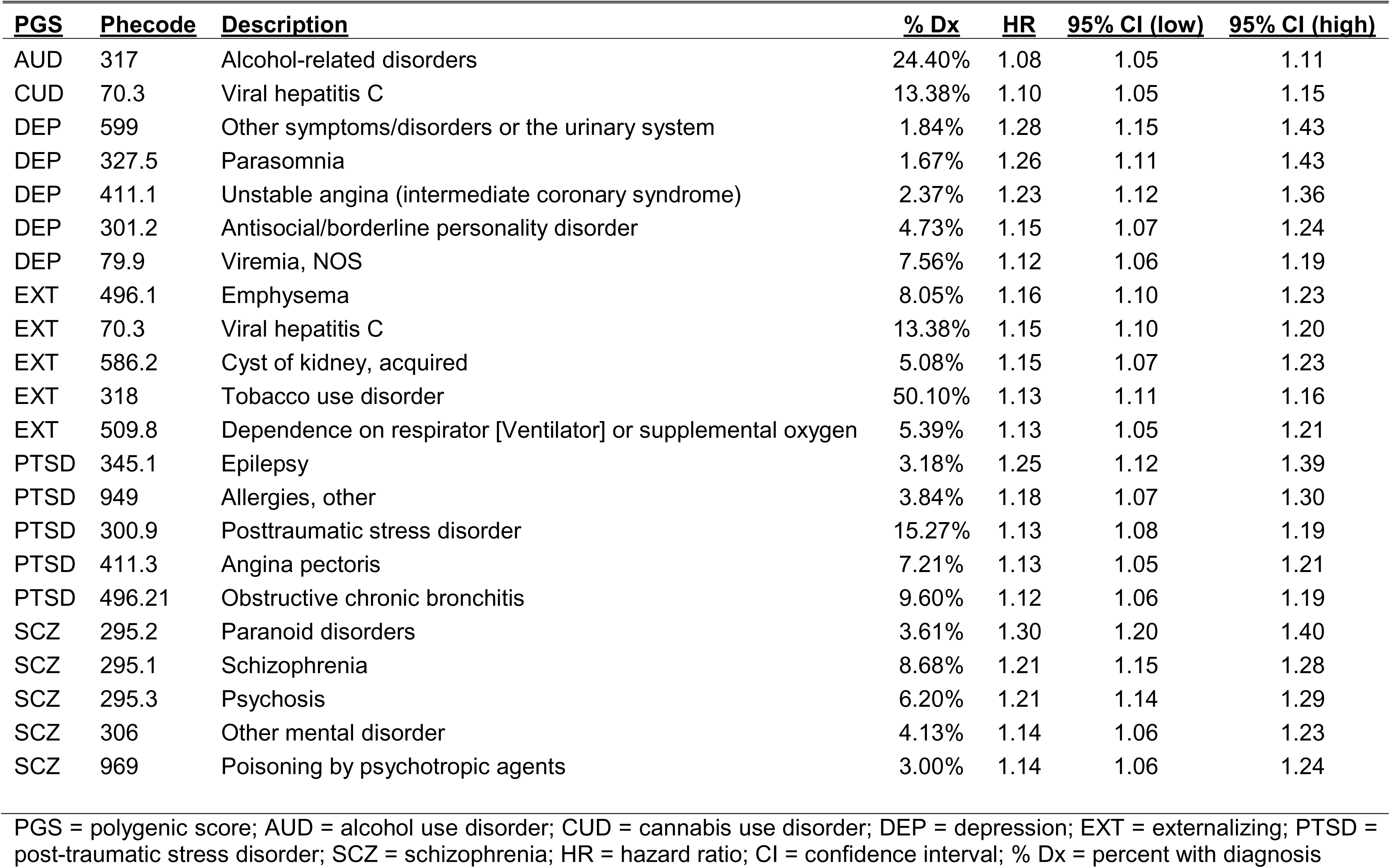
Selected Phenome Wide Significant Association.

PGSs were associated with multiple post-SUD-diagnosis comorbidities. For the SCZ PGS, the strongest associations were with SCZ-related diagnoses, including psychosis (HR = 1.21; 95% CI = 1.14, 1.28) and paranoid disorders (HR = 1.30; 95% CI = 1.20, 1.40). For PTSD the strongest associations were epilepsy (HR = 1.25; 95% CI = 1.12, 1.29) and cardiovascular conditions (angina pectoris: HR = 1.13; 95% CI = 1.05, 1.21). The EXT PGS was primarily related to potential consequences of substance use, such as smoking (emphysema: HR = 1.16, 95% CI = 1.10, 1.22; dependence on respirator or supplemental oxygen: HR = 1.13, 95% CI = 1.05, 1.21) or intravenous drug use (viral hepatitis C: HR = 1.15, 95% CI = 1.10, 1.20). The DEP PGS was primarily associated with subsequent psychiatric diagnoses (parasomnia: HR = 1.26, 95% CI = 1.11, 1.43; antisocial/borderline personality disorder: HR = 1.15, 95% CI = 1.07, 1.24). Lastly, the CUD PGS was only associated with subsequent viral hepatitis C infection (HR = 1.10, 95% CI = 1.05, 1.15).

Figure 3 presents two of the top associations between the PTSD and EXT PGSs and time to subsequent: 1) psychosis and 2) viral hepatitis C codes, respectively. Here PGSs were stratified by standard deviations (SD), ranging from −2 SD (red) to +2 SD (purple). While the overall effect of the PGSs was small, by the end of observation, the difference in probability of “survival” between these two extremes was approximately 12% lower for psychosis and 10% lower for viral hepatitis C in the +2 SD group. This sizeable difference reiterates how small effects can still have considerable effects when viewed over a longer time scale.

**Figure 3:**
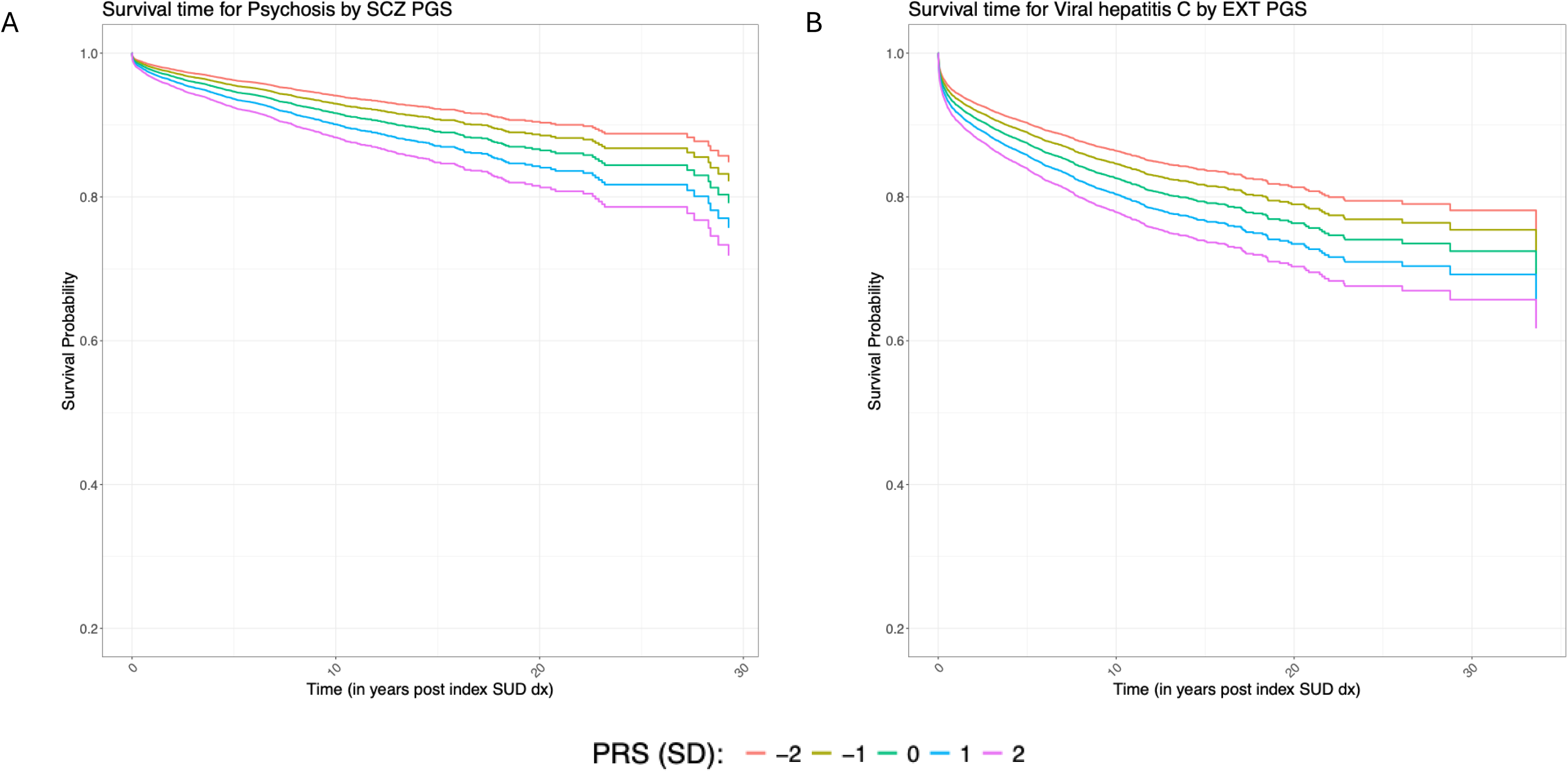
Survival plots for psychosis and viral hepatis C across SCZ and EXT PGS levels. Predicted survival curves for psychosis (Panel A) and viral hepatis C (Panel B) as a function of SCZ and EXT PGS levels, respectively (red = −2 SD, yellow = −1 SD, green = mean, blue = +1 SD, purple = +2 SD). All models adjusted for age at diagnosis, gender, genetic similarity, first 10 genetic principal components (PCS), pre-SUD comorbidity burden, and length of EHR.

Lastly, we repeated all of the previous analyses while including those with that only met criteria for tobacco use disorder (phecode 318) as part of our definitions of SUD diagnosis. This wider definition captured significantly more people (N = 67,729) and included 900 phecodes which occurred at 1% or more in the analytic sample. While there were additional significant associations within the survival PheWAS (162 phenome-wide significant associations, 18.11% of outcomes tested), the pattern of results largely mimicked those from the more restricted definitions in both sets of analyses. All of the results from the more expansive SUD definition are presented in the supplemental information (Supplemental Tables 5 – 8).

## DISCUSSION

Large-scale biobanks and genetic data are becoming increasingly prevalent in health research, offering the potential to explore multiple comorbidities simultaneously. The goal of precision medicine is harnessing these sources of information to personalize treatment and stratify risk. However, much of the work to date has focused on lifetime risk for a single diagnosis. In the current analyses, we expand upon existing research by focusing on risk longitudinally and within persons who have a SUD diagnosis to begin to understand what value risk factors might play in helping those who already have a diagnosis.

In terms of lifetime risk for SUD, we found multiple social and demographic factors associated with lifetime SUD diagnosis. These results largely mimic prior findings in All of Us (59) and in the population more broadly (51, 52), whereby those who largely occupy marginalized positions are at greater risk for having a SUD. The most striking finding is the gradient in income, whereby those reporting an annual household income less than $10k annually had ∼4 times the odds of SUD-diagnosis relative to those reporting annual incomes of $100k - $150k annually, reflecting either social determinants as major risk factors or the disabling impact of SUD on social mobility. The increased rate of diagnosis among those who identify as Black or African-American compared to Non-Hispanic Whites disappeared after accounting for other social determinants of health, suggesting the increased rates of diagnoses in this population may reflect socioeconomic inequalities and the role of systemic racism in limiting access to economic resources.

Focusing on genetic risk within persons who had a SUD diagnosis, 86 associations between PGS and subsequent phecodes remained significant after correcting for multiple testing (∼10% of outcomes tested), including associations between PGSs for SCZ, PTSD, and AUD and time to their corresponding diagnoses. These results increase confidence that associations are not merely spurious, as these PGSs demonstrated specificity for the expected outcomes. Additionally, these associations inform the degree to which PGS for psychiatric conditions may be relevant in those who already have a diagnosis for another psychiatric or substance use disorder. For example, PGS for both PTSD and SCZ were relevant for time to subsequent PTSD and schizophrenia diagnoses, respectively. Given the relatively strong genetic overlap between both PTSD and schizophrenia with SUDs (42, 48, 60), the continued relevance of these PGS within SUD cases demonstrates that having a diagnosis for a specific outcome (e.g. SUD) may not be sufficient to account for the genetic overlap between that trait and other diagnoses (e.g. PTSD or SCZ). Lastly, the association between the AUD PGS and subsequent AUD codes suggests that PGS not only capture lifetime risk for a diagnosis, but they may also capture aspects disease of severity, given the association with a repeated EHR codes, occurring earlier in those already diagnosed with SUD.

The survival PheWAS results also highlighted risk of comorbid conditions that occur after SUD diagnosis. A PGS for externalizing was associated with a variety of smoking-related conditions (e.g., chronic airway obstruction, use of a ventilator, etc.) as well as other potential consequences of SUD (e.g. viral hepatitis C). Prior work on externalizing PGS in both the Million Veteran Program and the Vanderbilt University Biobank revealed many of the same top associations (16, 34), helping add to the confidence in these associations. PGS for PTSD and depression were both associated with a variety of physical comorbidities, especially those related to cardiovascular disease and its risk factors. These associations are in line with broader epidemiological evidence which finds both depression and PTSD as consistent risk factors for the development of cardiovascular disease (61–63). SCZ PGS were most strongly associated with schizophrenia-related codes, including codes for paranoid disorders and psychosis, again consistent with other PheWAS of SCZ PGS (14, 15). Importantly, the SCZ PGS had the most significant associations, suggesting that while it is highly specific to schizophrenia risk, it captures other comorbidities when a diagnosis is not always available. Lastly, each of the polygenic scores were also associated with a *longer* time to a variety of diagnoses in the EHR. While this could be interpreted as some type of protective effect, the more likely explanation is those at greater risk are less likely to treatment (64, 65), especially as barriers to access, such as health insurance and economic resources, were key predictors in the social demographic correlates.

When we consider the results from the SDoH and PGS analyses together, we can see how the strong gradients across social conditions may be particularly relevant for considering *who* we are studying in EHR-based analyses of diagnosed cases. First, conditioning on a potential collider, such as SUD diagnosis, can bias downstream associations between PGS and outcomes in unpredictable ways (25, 66), so results should be interpreted with this potential bias in mind. Second, SDoH are clearly relevant for lifetime SUD risk and most likely important for associated comorbidities. Socioeconomic conditions may confound relationships between PGS and outcomes, since polygenic scores are confounded, at least in part, by environmental variance (67). While we cannot determine the degree to which socioeconomic may: 1) confound or 2) moderate the relationship between PGS and subsequent diagnoses, future analyses in All of Us that focus prospectively from time of enrollment can potentially better answer these questions.

This work has several important limitations. First, we lacked the social and demographic risk factors measured at time of diagnosis and could not establish the time ordering between these measures. As All of Us grows through recruitment, data generation, and passive data collection via constantly updating EHRs, we will be able to explore the prospective associations between important social determinants of health and diagnoses. Second, the use of EHR data, while convenient, is also confounded by access and healthcare utilization. We cannot discern whether associations are truly reflective of the progression in disease processes or whether they reflect propensity for help seeking when available in a healthcare setting. Third, this analysis focused on diagnoses that occurred after a recorded SUD. We did not address the scenario in which SUD codes follow other diagnoses (especially psychiatric diagnoses). However the framework we employed here could easily be extended to those other scenarios. Lastly, though we have attempted to be more inclusive in the genetic analyses, we still lacked sufficient GWAS summary statistics for other genetic similarity groups to include here. As a field, we must continue to strive to ensure results from genetic discoveries can be applied to all populations, equitably (56).

Harnessing the longitudinal data in the All of Us database, we have demonstrated the importance of various social conditions for the lifetime diagnosis of substance use disorders. Annual income is especially relevant in disparities of SUD. Additionally, we have also shown that in persons who have a SUD diagnosis, various measures of genetic risk were associated with time to subsequent diagnoses in their EHR, moving beyond typical cross-sectional approaches. While there is considerable work to be done for the use of social, clinical, and biological data within a healthcare setting, the current results demonstrate the potential of these approaches. Future work should endeavor to integrate across levels of analysis in a longitudinal framework.

## Data Availability

All data produced are available online via the All of Us Researcher Workbench.

## ACKNOWLEDGEMENTS

This study was supported in part by National Institute of Mental Health (R01MH125938: Drs Peterson, Bigdeli, Chatzinakos, and Meyers); the National Institute of Drug Abuse (R01DA050721: Dr Barr; R01DA060596: Drs Meyers, Barr, Chatzinakos, and Neale), and the National Institute on Alcohol Abuse and Alcoholism (R01AA030010: Drs Meyers, Barr, Chatzinakos, and Neale). Computing costs associated with this work was supported by R01MH125938 (PI: Peterson). The content of this article is solely the responsibility of the authors and does not necessarily represent the official views of the National Institutes of Health. The All of Us Research Program is supported by the National Institutes of Health, Office of the Director: Regional Medical Centers: 1 OT2 OD026549; 1 OT2 OD026554; 1 OT2 OD026557; 1 OT2 OD026556; 1 OT2 OD026550; 1 OT2 OD 026552; 1 OT2 OD026553; 1 OT2 OD026548; 1 OT2 OD026551; 1 OT2 OD026555; IAA #: AOD 16037; Federally Qualified Health Centers: HHSN 263201600085U; Data and Research Center: 5 U2C OD023196; Biobank: 1 U24 OD023121; The Participant Center: U24 OD023176; Participant Technology Systems Center: 1 U24 OD023163; Communications and Engagement: 3 OT2 OD023205; 3 OT2 OD023206; and Community Partners: 1 OT2 OD025277; 3 OT2 OD025315; 1 OT2 OD025337; 1 OT2 OD025276. In addition, the All of Us Research Program would not be possible without the partnership of its participants. We are grateful to Tadeusz H Wroblewski for sharing the image for survival models in All of Us.

